# Comparable seasonal pattern for COVID-19 and Flu-Like Illnesses

**DOI:** 10.1101/2021.02.28.21252625

**Authors:** Martijn J. Hoogeveen, Ellen K. Hoogeveen

## Abstract

**Background:** During the first wave of COVID-19 it was hypothesized that COVID-19 is subject to multi-wave seasonality, similar to Influenza-Like Illnesses since time immemorial. One year into the pandemic, we aimed to test the seasonality hypothesis for COVID-19.

**Methods:** We calculated the average annual time-series for Influenza-Like Illnesses based on incidence data from 2016 till 2019 in the Netherlands, and compared these with two COVID-19 time-series during 2020/2021 for the Netherlands. We plotted the time-series on a standardized logarithmic infection scale. Finally, we calculated correlation coefficients and used univariate regression analysis to estimate the strength of the association between the time-series of COVID-19 and Influenza-Like Illnesses.

**Results:** The time-series for COVID-19 and Influenza-Like Illnesses were strongly and highly significantly correlated. The COVID-19 peaks were all during flu season, and lows were all in the opposing period. Finally, COVID-19 meets the multi-wave characteristics of earlier flu-like pandemics, namely a short first wave at the tail-end of a flu season, and a longer and more intense second wave during the subsequent flu season.

**Conclusions:** We conclude that seasonal patterns of COVID-19 incidence and Influenza-Like Illnesses incidence are highly similar, in a country in the temperate climate zone, such as the Netherlands. Further, the COVID-19 pandemic satisfies the criteria of earlier respiratory pandemics, namely a first wave that is short-lived at the tail-end of flu season, and a second wave that is longer and more severe.

This seems to imply that the same factors that are driving the seasonality of Influenza-Like Illnesses are causing COVID-19 seasonality as well, such as solar radiation (UV), temperature, relative humidity, and subsequently seasonal allergens and allergies.

**HIGHLIGHTS:**

- Time-series analysis shows that COVID-19 and historic of Influenza-Like illnesses have highly similar seasonal patterns in the Netherlands.
- COVID-19 satisfies the criteria of earlier flu-like pandemics, namely a short cycle at the end of flu season, and a longer, more intense cycle during the subsequent flu season.
- The implication is that the seasonal factors driving flu season, are also responsible for COVID-19 seasonality.
- We developed and applied a new method to determine seasonality, encompassing comparative time-series analysis, a standardized logarithmic infection scale, and qualitative seasonality criteria.

## 1. INTRODUCTION

During the first wave of COVID-19 it was hypothesized that COVID-19 is subject to multi-wave seasonality [1, 2], comparable to other respiratory viral infections and pandemics since time immemorial [3, 4]. It is observed that the COVID-19 community outbreaks have a similar pattern as other seasonal respiratory viruses [5, 6, 7]. Already during the first COVID-19 cycle the data suggested seasonality, comparable to the seasonality of Influenza-Like Illnesses (ILI), although the time-series were typically too short for definitive conclusions [8]. Currently, we are one year into the COVID-19 pandemic, and we can witness in the temperate climate zone in the Northern Hemisphere, a second wave that appears to rise and peak during the boundaries of a typical flu season, as the first cycle before.

Until now, it is not yet confirmed that COVID-19 behaves as seasonal as ILI. Therefore, we aim to test our hypothesis that COVID-19 has a similar seasonal pattern as ILI in a country in the temperate climate zone as the Netherlands. To test our hypothesis, we performed time-series analysis to compare the COVID-19 cycles with the multi-wave seasonality patterns of flu-like illnesses. In addition, we analyzed to what degree the COVID-19 pandemic fulfills the qualitative characteristics of earlier flu-like pandemics and seasonality as mentioned by Fox et al. Particularly, a short first wave at the tail end of a flu season, and a more severe second wave during the following flu season. We further expect peaks to occur within the seasonal boundaries between week 33 (± 2 weeks) and week 10 (± 5 weeks), and the nadir in the opposing period which coincides with the allergy season [8, 9].

The main objective of this study is to provide a predictive model for subsequent COVID-19 seasonal cycles.

## 2. METHODS

### 2.1 Data

#### Incidence of Influenza-Like Illnesses

We used data from the Dutch State Institute for Public Health (RIVM) gathered by the Dutch institute for research of the health care (Nivel) about weekly flu-like incidence (WHO code “ILI” - Influenza-Like Illnesses). ILI is defined by the WHO as a combination of a measured fever of ≥ 38°C, and a cough, with an onset within the last 10 days. The Dutch ILI reports are gathered from primary medical care. Primary medical care is the first-line healthcare provided by general practitioners to their registered patients as typical in the Netherlands, with its current population of 17.4 million. The reports are confirmed by a positive RIVM laboratory test for ILI [10].

The flu-like incidence metric is a weekly average based on a representative group of 40 primary care units. It is calculated using the number of influenza-like reports per primary care unit divided by the number of patients registered at that unit. This is then averaged for all primary care units in the Netherlands, extrapolated to the entire population, and reported as the ‘ILI incidence per 100,000 citizens in the Netherlands’. The datasets run from week 1 of 2016 up to week 52 of 2019 to preclude the COVID-19 pandemic and avoid the impact of lockdowns and other COVID-19 related restrictions on ILI in the 2020/2021 season. We used these data to calculate the average ILI incidence per week (n = 52) as our baseline times series.

#### COVID-19 incidence

To calculate the COVID-19 incidence, we used the RIVM data set which reports the daily COVID-19 incidence per municipality [11]. The incidence per municipality is based on positive COVID-19 tests that are reported via the local municipality health services (Gemeentelijke Gezondheidsdienst; GGD), that are under the control of the RIVM. We aggregated the crude number into a weekly COVID-19 incidence for the Netherlands per 100,000 citizens, to create a metric on the same scale as the standard ILI metric. We calculated the values from week 13, 2020, the peak of the short first COVID-19 cycle in the Netherlands, till week 5, 2021 (n = 45). We assume that the cycles themselves are sufficiently representative for time-series analysis, even though the COVID-19 incidence during the first cycle is most likely underestimated compared to incidence during the second cycle, due to test bias. With test bias, we mean that both the method of testing and the test capacity, altered during the development of the COVID-19 pandemic. Especially, at the start of the pandemic, there was a shortage of test capacity in the Netherlands. The capacity shortage had considerably improved since the end of March, i.e., week 13 of 2020, but only in the first months of 2021 reached sufficient levels.

Therefore, as a sensitivity analysis, we used a second dataset from RIVM, which is based on data from the Dutch national intensive care evaluation foundation [12]. Based on hospital admissions, the RIVM estimated the COVID-19 incidence in the Netherlands, assuming a delay of at least 7 days between the COVID-19 infection and hospital admission. This dataset provides the average COVID-19 incidence per day with a 95% confidence interval (95% CI). Again, we calculated the average weekly COVID-19 incidence for the Netherlands per 100,000 citizens, to create a metric on the same scale as the standard ILI metric.

### 2.2 Statistical analysis

Variables are presented with their means (M) and standard deviations (SD). We calculated correlation coefficients to test the hypotheses and to assess the strength and direction of relationships.

Because the time-series were nonlinear and somewhat skewed, we used the log10 transformation before applying linear regression and calculating correlation coefficients, which requires a normal distribution. After the log10 transformation, we multiplied the data with factor 2 to create an intuitive scale from 0 to 10 for ILI and COVID-19, which is comparable to the 1 to 12 logarithmic Richter scale for earthquakes [13]. The logarithmic scale we elaborated is rational and plots exponential characteristics on a linear infections scale [14]. We added descriptive labels to each scale as an aid for qualitative interpretation in intuitive, layman terms (see Table 1). Other advantages are that it makes a comparison between different epidemics or pandemics, and thus external validation, easier. Finally, it enlarges the critical early stages of an epidemic, and it reduces the extreme peaks and resulting test bias because of test capacity overloads.

**Table 1:** Logarithmic Infection Scale.

Logarithmic scale [1, 10] of Influenza-Like Illnesses or COVID-19 (or other) incidence because of the exponential nature of epidemics, with proposed qualitative descriptions for convenience.
| $2 \times \text{LOG}_{10}(\text{Incidence}/100\text{K})$ | Incidence<br>(/100K citizens) | Incidence (% of<br>population<br>infected) | Description |
| --- | --- | --- | --- |
| 0 | 1 | 0.001% | Isolated incidents |
| 1 | 3 | 0.003% | Mild outbreak |
| 2 | 10 | 0.01% | Moderate outbreak |
| 3 | 30 | 0.03% | Severe outbreak |
| 3.5 | 58 | 0.06% | Epidemic threshold (example) |
| 4 | 100 | 0.1% | Mild Epidemic |
| 5 | 300 | 0.3% | Moderate epidemic |
| 6 | 1000 | 1% | Severe epidemic |
| 7 | 3000 | 3% | Very severe epidemic |
| 8 | 10000 | 10% | Public health catastrophe |
| 9 | 30000 | 30% | Severe public health catastrophe |
| 10 | 100000 | 100% | Total public health catastrophe |

Linear regression (F-test) on the ILI and COVID-19 time-series is performed as a sensitivity analysis and used *descriptively* to determine the strength of the relationship between the COVID-19 and ILI time-series. More in detail to determine the equation using estimates and intercept values, probability, significance level, F-value, and the Multiple R squared correlation to understand the predictive power of the respective relation. Standard deviations and errors and degrees of freedom (DF) were used as input for calculating the 95% probability interval.

We have reported the results in APA style, adapted to journal requirements.

Correlations are calculated manually in Excel, and for linear regression Graphpad 2021 is used (which we benchmarked on R version 3.5).

## 3. RESULTS

### 3.1 Data analysis

The means and standard deviations of the dataset are summarized in Table 2.

**Table 2:**
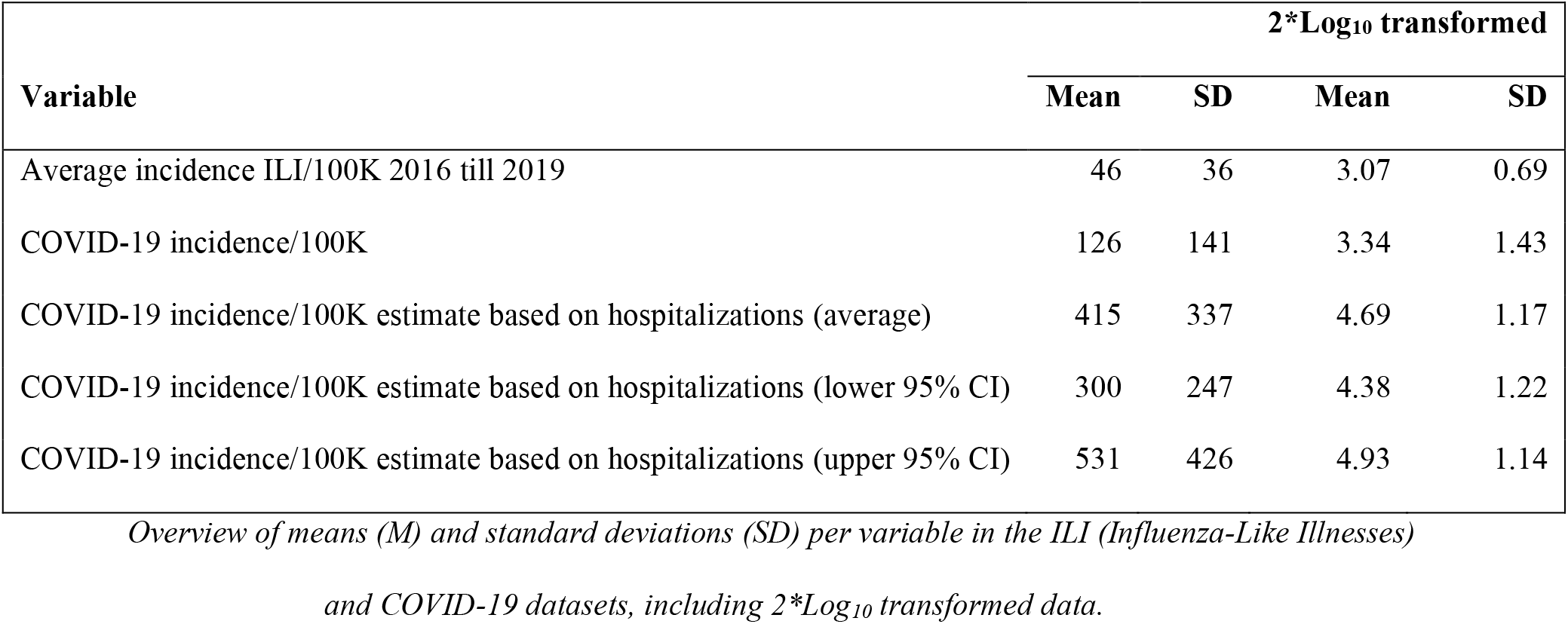
overview means (M) and standard deviations (SDs)

Figure 1 shows a short first COVID-19 wave at the tail end of the 2019/2020 flu season, and a more severe second wave during the 2020/2021 flu season in terms of total incidence. The peaks are all within the seasonal boundaries between week 33 (± 2 weeks) and week 10 (± 5 weeks), and the nadirs in the opposing period. The hospitalizations-based estimates for COVID-19 incidence provide likely a more realistic picture of especially, the first wave, given test bias. However, on a logarithmic scale, the first COVID-19 wave appears more visually comparable in both time-series.

**Fig. 1.**
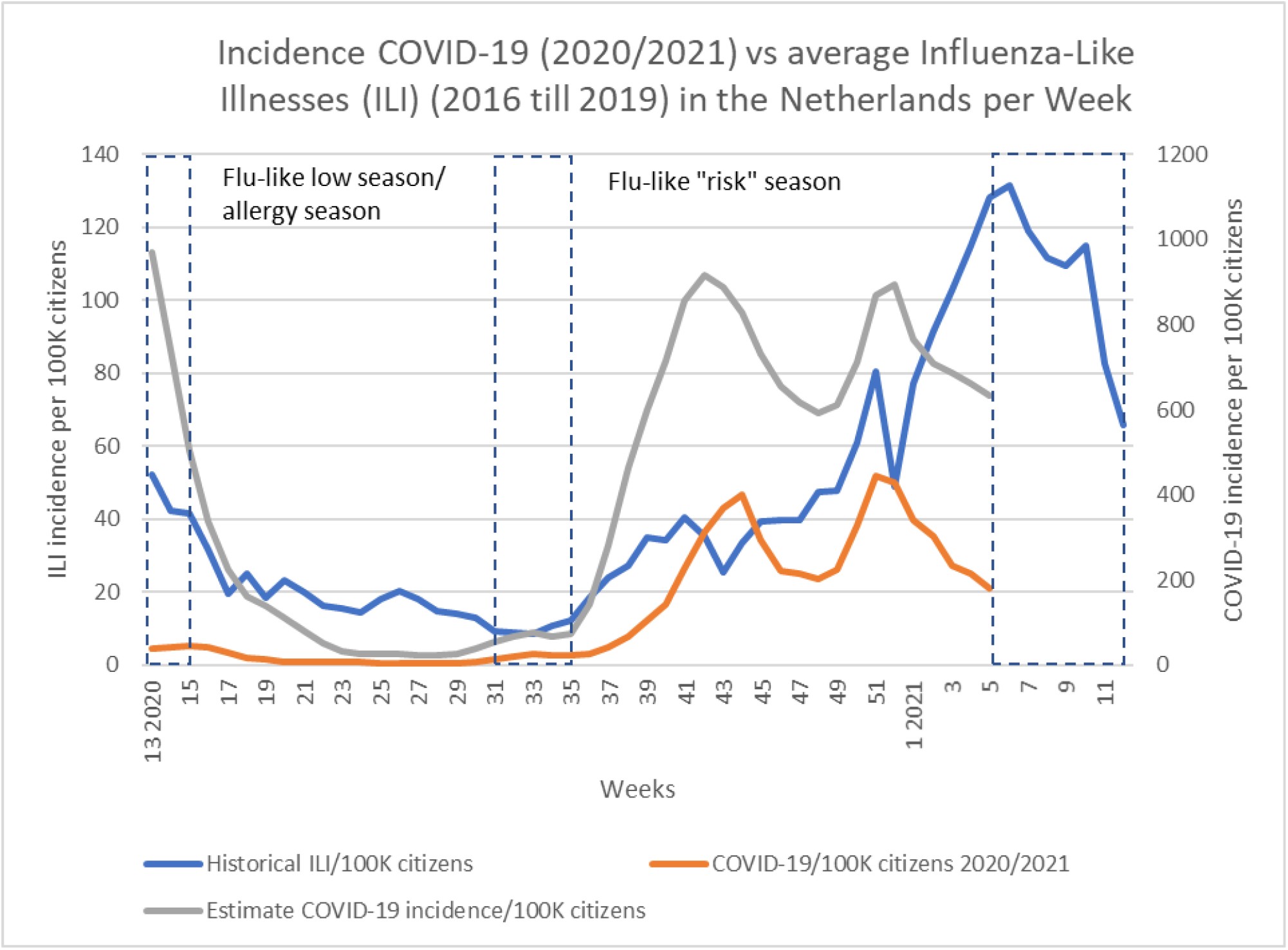
Historical ILI (Influenza-Like Illnesses) incidence (2016 till 2019) versus COVID-19 incidence per 100K citizens during the 2020/2021 season. Peaks are all during flu season, and lows during the opposite season. The shaded periods are the typical period in which seasonal switching occurs.

On our logarithmic infection scale (Fig. 2), the estimated COVID-19 incidence tops around 6 (severe epidemic level), and the nadir bottoms out around 3 (severe outbreak level). Interestingly, on this scale, it becomes visible that COVID-19 incidence starts to rise slightly earlier than what is usual for ILI (week 33 ± 2 weeks).

**Fig. 2.**
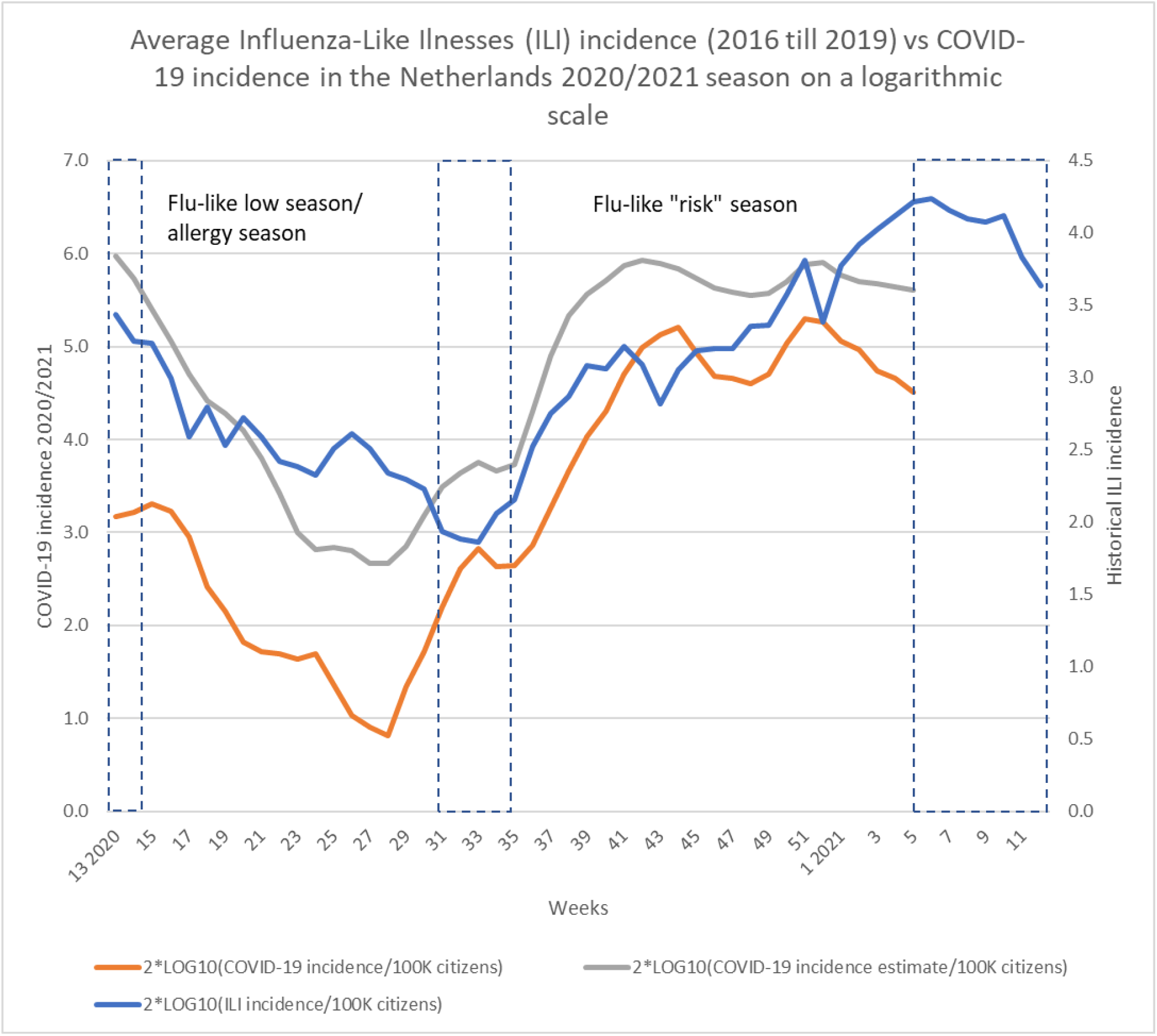
Historical ILI (Influenza-Like Illnesses) incidence (2016 till 2019) versus COVID-19 incidence per 100K citizens (2020/2021) on the 1 to 10 logarithmic scale. This figure visualizes the similarity of the COVID-19 time-series based on hospitalizations with the historic ILI time-series. The shaded areas are the typical seasonal switching periods.

### 3.2 Statistical outcomes

The COVID-19 time-series strongly and highly significantly correlates with the ILI time-series *r*(45) = 0.75 (p < 0.00001). The hospitalizations-based, COVID-19 time-series, providing estimates that control for test bias, correlated even somewhat stronger, (*r*(45) = 0.798, p < 0.00001), and as significantly (Fig. 3). The correlations (95% CI) of the estimated COVID-19 incidence time-series are almost equal, respectively *r*(45) = 0.788, p < 0.00001 and *r*(45) = 0.803, p < 0.00001. Therefore, we conclude that the COVID-19 time-series have a similar wave pattern to the ILI time-series, which have long been established as being seasonal. Furthermore, the COVID-19 peaks, similar to ILI peaks, all occur during flu season, i.e., between week 33 (± 2 weeks) and week 10 (± 5 weeks). In addition, the COVID-19 nadirs, similar to ILI nadirs, occur all in the opposing allergy season.

**Fig. 3.**
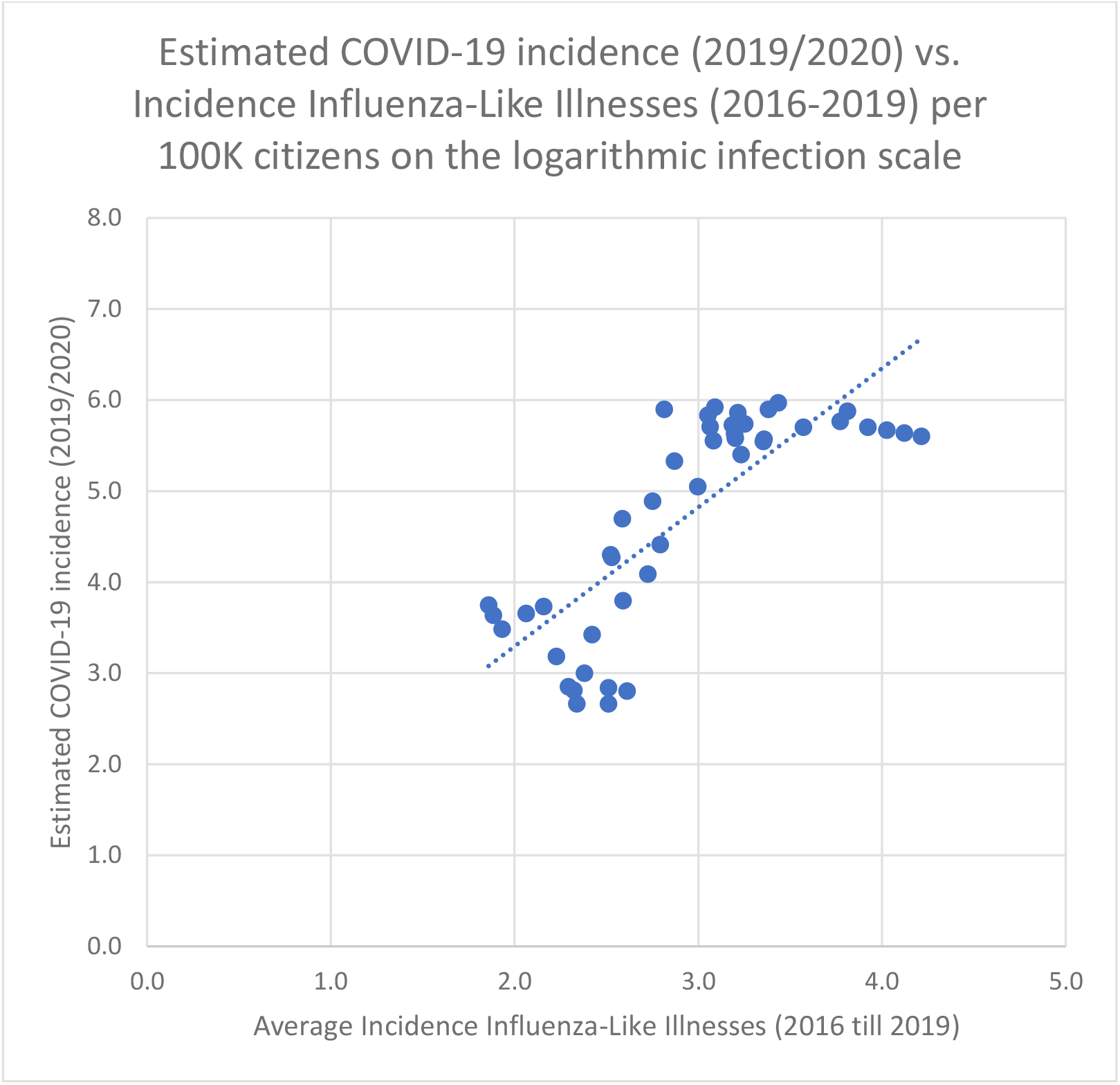
Scatter diagram showing the relation between the estimated COVID-19 incidence/100K citizens and the seasonal, average incidence of Influenza-Like Illnesses (ILI)/100K citizens in the Netherlands of the preceding 4 years.

As a second sensitivity analysis, we performed univariate regression analyses between both the COVID-19 time-series, and the average ILI time-series. The outcomes were again highly significant: respectively *F*(1, 43) = 61.45, p < 0.0001, and *F*(1, 43) = 81.18, p < 0.0001 and the correlations (*r*^*2*^) are moderate to strong (see Table 3).

**Table 3:**
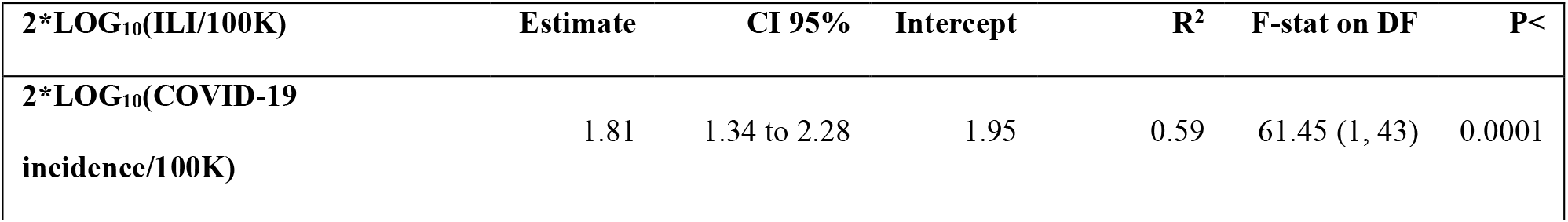
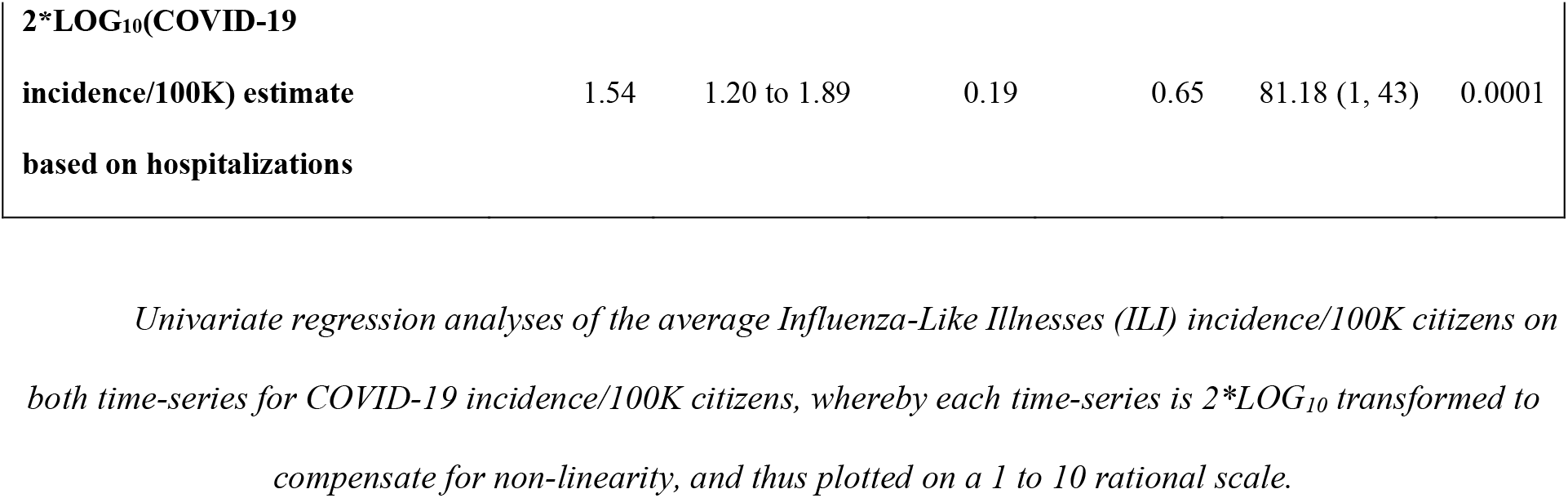
outcomes linear regression.

## 4. DISCUSSION

Given the strong, and highly significant, correlations between the ILI and COVID-19 time-series, we conclude that COVID-19 incidence follows a seasonal pattern that is similar to ILI incidence in a country in the temperate climate zone, such as the Netherlands. Moreover, the COVID-19 peaks are all during flu season, and lows are all in the opposing period as expected. Furthermore, the COVID-19 time-series is in accordance with the two characteristics of earlier pandemics [4], namely a short first wave at the tail-end of a flu season, and a longer and more intense second wave during the subsequent flu season. This implies that the subsequent endings and starts of each following wave are more or less predictable. If the history of pandemics is followed, the third COVID-19 wave would be less severe than the second one. Though nowadays vaccination will be a more important factor in determining the amplitude of the subsequent waves.

Interestingly, all over Europe, the COVID-19 cycles were all more or less in sync with the Dutch COVID-19 cycle [15], and thus ILI seasonality, independent of the start of the first cycle, the severity of lockdown measures taken, and given that herd immunity is not yet reached. The seasonality pattern of COVID-19 appears to be influenced though not caused by social distancing and lockdown measures as these measures were mainly anti-cyclical and following the trend. They were increasingly applied to flatten the curve after COVID-19 incidence increases, gradually lifted after the sharper than expected COVID-19 downcycles in Spring and Summer, and only re-applied after the second wave seriously kicked in, during Autumn and Winter. It is beyond our research to quantify the considerable impact of lockdown and social distancing measures, although it might explain that COVID-19 incidence on the logarithmic scale (see Fig. 2) starts to rise slightly earlier than what is usual for ILI (week 33 ± 2 weeks) as social distancing and lockdown measures were increasingly relaxed and ignored in this period.

What environmental factors have caused COVID-19 seasonality? We have analyzed before that the likely inhibiting factor causing ILI seasonality, before or during COVID-19, are seasonal allergens (i.e, pollens) and seasonal allergies [8, 9], given that meteorological factors alone are not sufficient to explain the seasonality of ILI [16] or COVID-19 [17]. We used data from a pollen station in Helmond, the Netherlands (latitude 51.48167, longitude 5.66111). On the other hand, we identified solar radiation (UV) as an ILI/COVID-19 co-inhibitor, and it is well-established that dry, warm, and sunny weather stimulates the maturation and dispersion of pollens.

The inverse seasonality of seasonal allergens and ILI including COVID-19 is independently confirmed by a recent Chicago (latitude 41.85003, longitude −87.65005) study that covered not only pollens but also mold spores [18]. These findings seemingly contradict an early international study of pollen and COVID-19 [19], which suffers from the same issues as many other early environmental COVID-19 studies: both sub-seasonal bias and COVID-19 test bias. Sub-seasonal bias, means here that a too limited time-series sample is used, namely from January till the beginning of April 2020, which coincides with only a small part of the early upswing in COVID-19 incidence in most countries, and mainly the upswing of allergens in the northern hemisphere. Further, the assumed lag of precisely 4 days seems to be problematic as people typically do not get immediately tested when they show the first COVID-19 symptoms. Additionally, conflicting local outcomes were selectively removed from the study. And, finally, in this short time window used it might have been better to correlate with a sound R0 estimate, correcting for test bias, than only the raw incidence figures, to be able to discern deceleration/acceleration effects during the upswing.

We think that studies that are more geographically focused, and work with longer time-series, are in the current phase more meaningful for analyzing the pollen effect on COVID-19 incidence while controlling for deviations in other local circumstances. Therefore, we would expect that, ceteris paribus, the results of our COVID-19 and ILI pattern comparison in this study, implies that a 12-month time-series analysis of the R0 of COVID-19 and pollen concentrations would falsify the outcomes of this international study, at least for the Netherlands.

Although the allergenic role of pollen is widely known [20, 21, 22], its role in immuno-activation [23] is highlighted only recently. Owing to extensive COVID-19 research, pathophysiological explanations have been established upon the observation that allergic diseases are associated with lower rates of COVID-19 hospitalizations [24, 25]. An explanation is provided by Jackson et al. [26], who proved that allergic sensitization and allergen natural exposure are inversely related to membrane-bound angiotensin-converting enzyme 2 (ACE-2) expression, whereby it is known that severe acute respiratory syndrome coronavirus 2 (SARS-CoV-2) uses the ACE-2 receptor to gain cell entry, leading to COVID-19 [27]. Additionally, it is hypothesized that seasonal allergens, especially pollen and mold spores, compete with flu-like viruses for access to another receptor, the Toll-Like Receptor 4 (TLR4) [18], in an attempt to explain the inverse seasonality of seasonal allergens and flu-like illnesses, both for people with and without allergic diseases.

Another explanation is based on the higher eosinophil count in children with allergic diseases than COVID-19 patients [28], whereby eosinophils are known to clear viral load, and contribute to the recovery from viral infections, supposedly including COVID-19 [29].

Furthermore, histamine and IgE serum levels are elevated in allergic rhinitis and other atopy patients which downplay other anti-viral responses (plasmacytoid dendritic cells and interferon-α) but might thus prevent the cytokine storm and hyper-inflammation that typically mark severe outcomes of respiratory diseases, including COVID-19 [30] and influenza. And, it is discussed that homologies between SARS-CoV-2 and allergen proteins, especially from the spores of molds (Aspergillus fumigatus) and the pollen of grasses (Phleum pratense) may direct T cell-mediated heterologous immune responses [31] which might help explain the protective effect of allergies against COVID-19 hospitalization as well.

Finally, it is well-established that pollens have anti-viral phytochemicals [32], which is also the case for mold spores [33]. Although the nature of the bioaerosol interaction with COVID-19 viruses is not well investigated yet it can be imagined that pollens and spores might compete with viral bioaerosol in the environment.

### Methodological concerns

Test bias, especially for new viruses such as COVID-19, is a major methodological challenge. The approach to use more reliable metrics like the number of hospitalizations to generate an alternative incidence metric appears to be a good method to control for test bias. We could observe that the test bias slightly reduced the strength of correlations, which in our case did not affect the conclusions. During 2020-2021, the test capacity was scaled up significantly and sufficiently, which reduced the test bias concern over time.

Another sound approach used seems to be excess mortality estimates. In this study, we decided not to use the latter, as there are other known factors than seasonal viruses that cause excess mortality, such as the heatwave during the Summer of 2020. Thus, using excess mortality would just introduce a measurement validity concern.

## 5. CONCLUSION

The COVID-19 pandemic in the Netherlands is till now as seasonal as flu-like illnesses given the highly significant and strong correlations between both time-series. But, also given that COVID-19 waves till now rise between the temporal boundaries (week 10 ±5 weeks and week 33 ±2 weeks) of the typical flu-like season in The Netherlands, and go down in the opposing periods. Further, the COVID-19 pandemic satisfies the qualitative criteria of earlier respiratory pandemics since 1889: the first wave is short-lived at the tail-end of flu season, the second wave is longer and more severe, peaks fall within the boundaries of flu-like season, and the lows are during the boundaries of the opposing season.

This logically seems to imply that, ceteris paribus, the same factors that are driving the seasonality of Influenza-Like Illnesses are causing COVID-19 seasonality, such as solar radiation (UV), temperature, relative humidity, and subsequently seasonal allergens and allergies.

## Data Availability

Data is fully based on publicly available sources.

https://data.rivm.nl/covid-19/COVID-19_prevalentie.json

https://www.rivm.nl/griep-griepprik/feiten-en-cijfers

https://data.rivm.nl/geonetwork/srv/dut/catalog.search#/metadata/1c0fcd57-1102-4620-9cfa-441e93ea5604

## FOOTNOTES PAGE

**Martijn Hoogeveen:** Conceptualization, Methodology, Data Curation, Formal Analysis, Writing, Investigation, Visualization, Resources

**Ellen Hoogeveen:** Methodology, Reviewing and Editing, Resources

## Funding

This research did not receive any specific grant from funding agencies in the public, commercial, or not-for-profit sectors.

## Declarations of interest

None.

## Data statement

The links to the COVID-19 datasets are provided in the reference list and a reference to the source of Influenza-Like Illnesses. Upon request, the data used for this manuscript is available for inspection, but for other purposes we kindly refer to the respective copyright-holder(s).

## MATERIALS

### Linear Regression

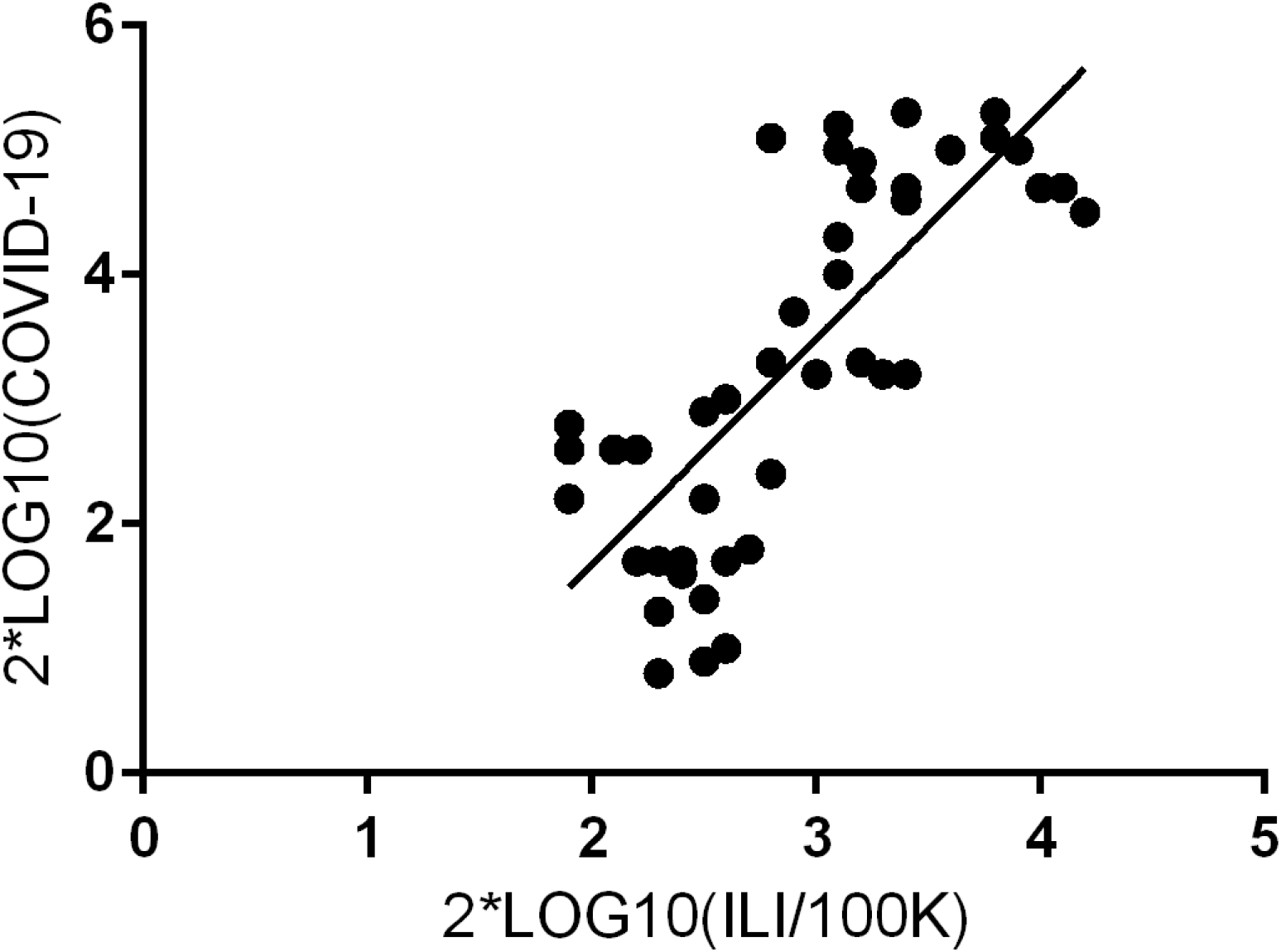

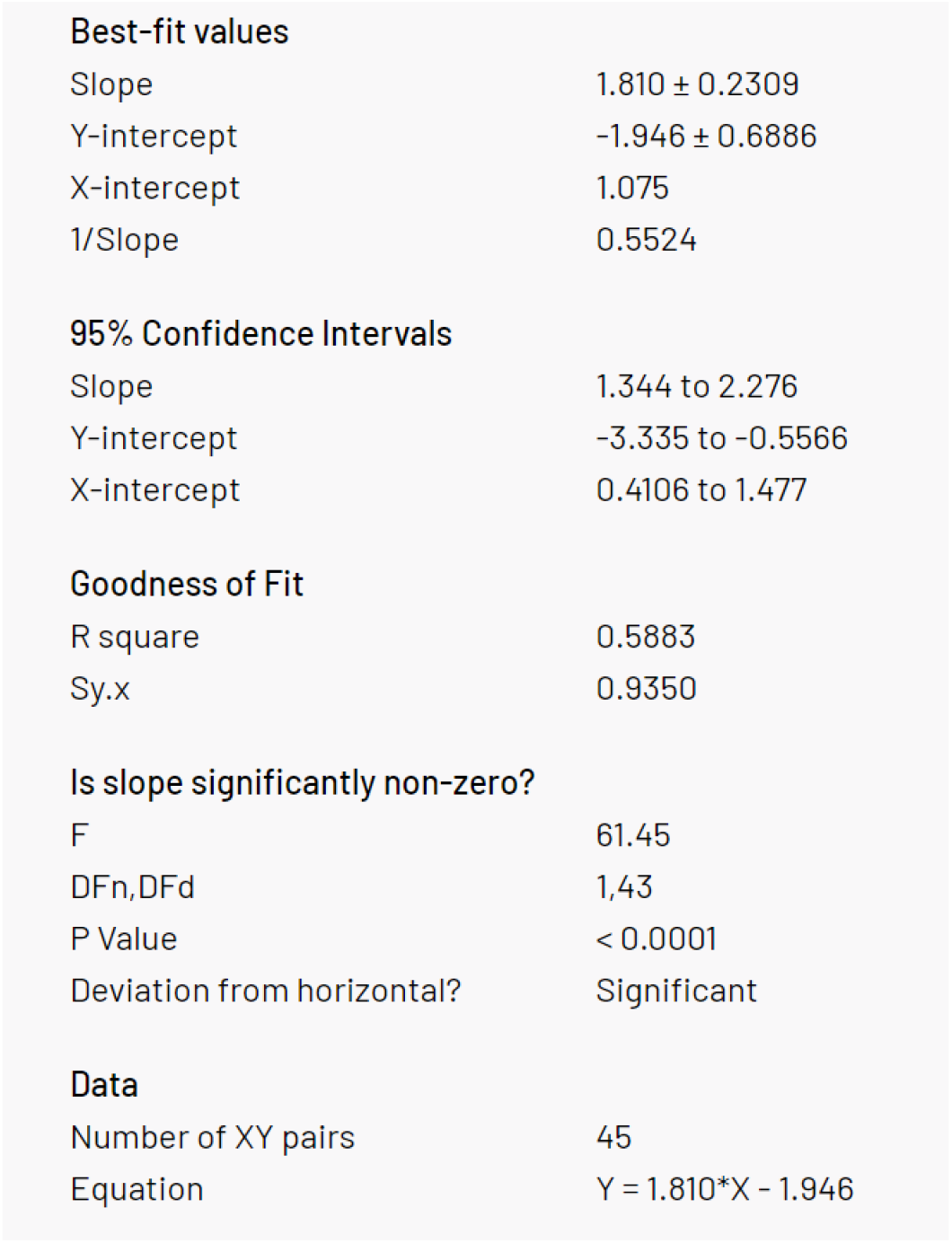

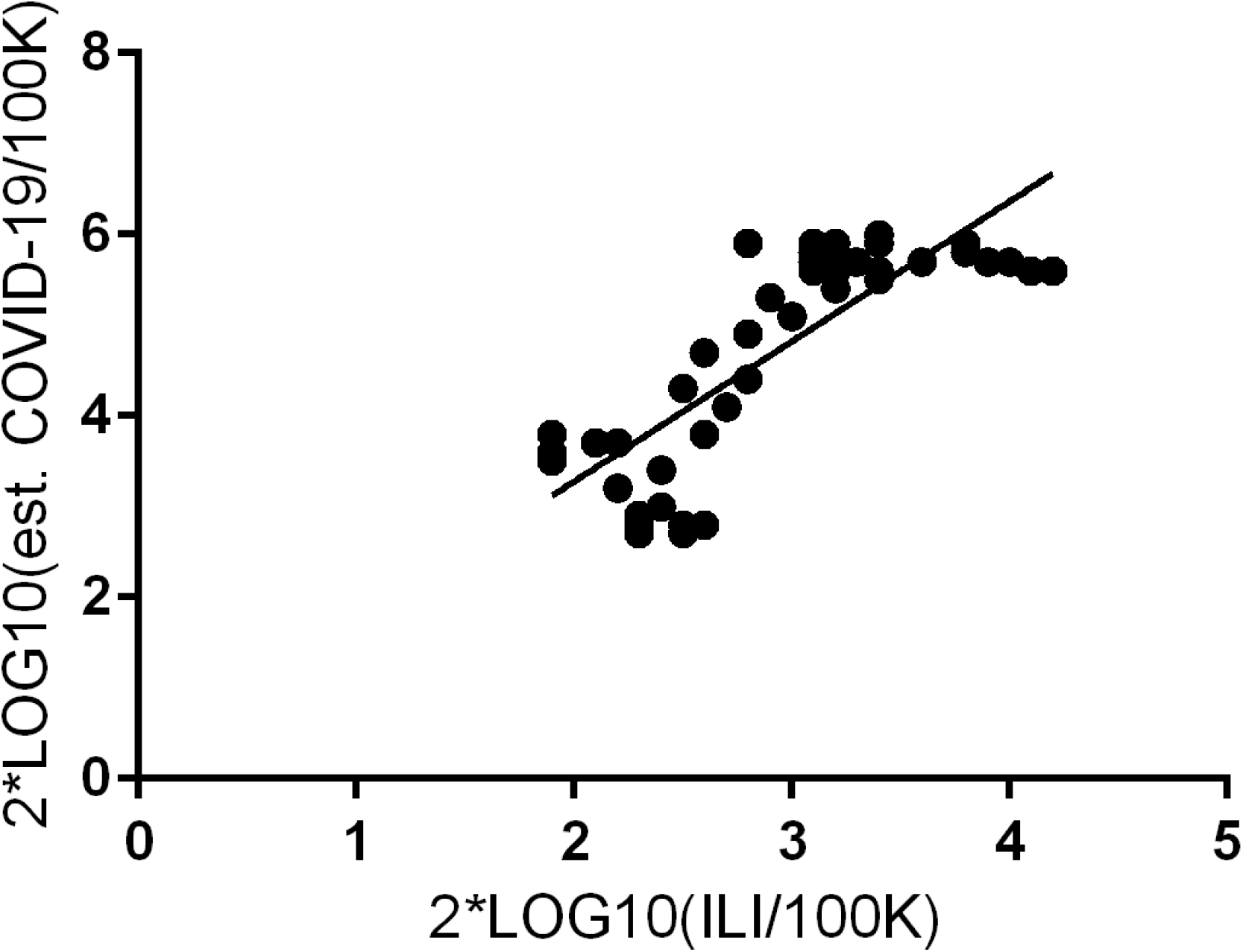

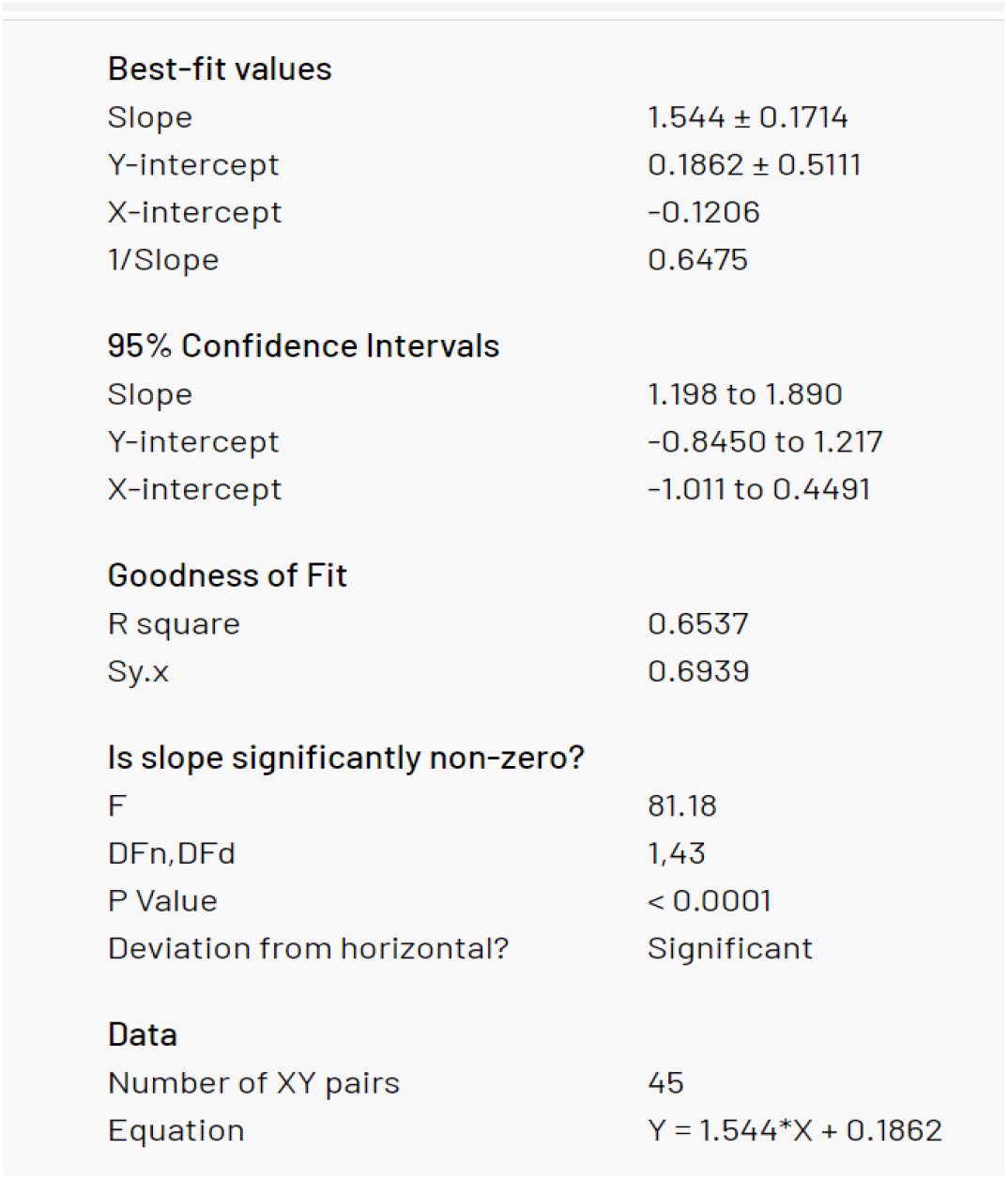

## Notes

### Competing Interest Statement

The authors have declared no competing interest.

### Summary of Updates

Improved the images, corrected grammar and a few minor typos, and applied AMA referencing style.

## REFERENCES

1. Kissler SM, Tedijanto C, Goldstein E, Grad YH, Lipsitch M. Projecting the transmission dynamics of SARS-CoV-2 through the post-pandemic period. Science. 2020;368(6493): 860–868, https://doi.org/10.1126/science.abb5793.

2. Grech V, Cuschieri S, Gauci C. COVID-19: The possible seasonal shape of things to come. Early Hum Dev. 2020 Nov 12: 105262, https://doi.org/10.1016/j.earlhumdev.2020.105262.

3. Moriyama M, Hugentobler WJ, Iwasaki A. Seasonality of Respiratory Viral Infections. Annual Review of Virology. 2020;7:1,https://doi.org/10.1146/annurev-virology-012420-022445.

4. Fox SJ, Miller JC, Meyers LA. Seasonality in risk of pandemic influenza emergence. PLoS Comput Biol 2017; 13(10): e1005749, https://doi.org/10.1371/journal.pcbi.1005749.

5. Sajadi MM, Habibzadeh P, Vintzileos A, Shokouhi S, Miralles-Wilhelm F, Amoroso A. Temperature, Humidity and Latitude Analysis to Predict Potential Spread and Seasonality for COVID-19. 2020 March 5, https://doi.org/10.2139/ssrn.3550308.

6. Poole L. Seasonal Influences On The Spread Of SARS-CoV-2 (COVID19), Causality, and Forecastabililty (3-15-2020) (March 15, 2020), https://doi.org/10.2139/ssrn.3554746.

7. Burra P, Soto-Díaz K, Chalen I, Gonzalez-Ricon RJ, Istanto D, Caetano-Anollés G. Temperature and Latitude Correlate with SARS-CoV-2 Epidemiological Variables but not with Genomic Change Worldwide. Evolutionary Bioinformatics. 2021 January, https://doi.org/10.1177/1176934321989695.

8. Hoogeveen MJ, Van Gorp Ecm, Hoogeveen EK. Can pollen explain the seasonality of flu-like illnesses in the Netherlands? Sci Total Environ. 2021 Feb 10;755(Pt 2):143182, https://doi.org/10.1016/j.scitotenv.2020.143182.

9. Hoogeveen MJ. Pollen likely seasonal factor in inhibiting flu-like epidemics. A Dutch study into the inverse relation between pollen counts, hay fever and flu-like incidence 2016–2019, Sci Total Environ. 2020 July 20; 727:138543, https://doi.org/10.1016/j.scitotenv.2020.138543.

10. RIVM/ILI data. Griep en griepprik. Feiten en cijfers. https://www.rivm.nl/griep-griepprik/feiten-en-cijfers. Accessed May 15, 2020.

11. [dataset] RIVM/GGD. COVID-19 incidence per Dutch municipality by GGDs. https://data.rivm.nl/geonetwork/srv/dut/catalog.search#/metadata/1c0fcd57-1102-4620-9cfa-441e93ea5604. Accessed February 14, 2021.

12. [dataset] RIVM/NICE. Estimated prevalence COVID-19 on the basis of a 95% confidence interval. https://data.rivm.nl/covid-19/COVID-19_prevalentie.json. Accessed February 14, 2021.

13. Boore DM. The Richter scale: its development and use for determining earthquake source parameters, Tectonophysics. 1989;166(1–3): 1–14, https://doi.org/10.1016/0040-1951(89)90200-X.

14. Steffen R. Travel vaccine preventable diseases—updated logarithmic scale with monthly incidence rates, Journal of Travel Medicine. 2018;25(1): tay046, https://doi.org/10.1093/jtm/tay046.

15. Reuters COVID-19 tracker Europe https://graphics.reuters.com/world-coronavirus-tracker-and-maps/regions/europe/. Accessed on February 28, 2021.

16. Tamerius J, Nelson MI, Zhou SZ, Viboud C, Miller MA, Alonso WJ. (2011). Global influenza seasonality: reconciling patterns across temperate and tropical regions. Environmental health perspectives, 119(4), 439–445, https://doi.org/10.1289/ehp.1002383.

17. Kerr GH, Badr HS, Gardner LM, Perez-Saez J, Zaitchik BF. Associations between meteorology and COVID-19 in early studies: Inconsistencies, uncertainties, and recommendations. One Health. 2021;12:100225, https://doi.org/10.1016/j.onehlt.2021.100225.

18. Shah RB, Shah RD, Retzinger DG, Retzinger AC, Retzinger DA, Retzinger GS. Confirmation of an Inverse Relationship between Bioaerosol Count and Influenza-like Illnesses, Including COVID-19. On the Contribution of Mold Spores. MedrXiv 21251322 [preprint]. Feb 16, 2021 [cited March 1, 2021]. Available on: https://doi.org/10.1101/2021.02.07.21251322.

19. Damialis A, Gilles S, Sofiev M, Sofieva V, Kolek F, Bayr D, Plaza MP, Leier-Wirtz V, Kaschuba S, Ziska LH, Bielory L, Makra L, Del Mar Trigo M, COVID-19/POLLEN study group, Claudia Traidl-Hoffmann. Higher airborne pollen concentrations correlated with increased SARS-CoV-2 infection rates, as evidenced from 31 countries across the globe. Proceedings of the National Academy of Sciences. 2021 Mar;118(12): e2019034118, https://doi.org/10.1073/pnas.2019034118.

20. Klemens C, Rasp G, Jund F, Hilgert E, Devens C, Pfrogner E, Kramer MF. Mediators and cytokines in allergic and viral-triggered rhinitis. Allergy & Asthma Proceedings. 2007;28(4): 434–441.

21. Rosenwasser LJ. Current Understanding of the Pathophysiology of Allergic Rhinitis. Immunology and Allergy Clinics. 2011;31(3): 433–439.

22. Howarth PH. Allergic rhinitis: not purely a histamine-related disease. European Journal of Allergy and Clinical Immunology 2000;55(64): 7–16.

23. Brandelius A, Andersson M, Uller L. Topical dsRNA challenges may induce overexpression of airway antiviral cytokines in symptomatic allergic disease. A pilot in vivo study in nasal airways. Respiratory Medicine. 2020; 108(12): 1816–1819, https://doi.org/10.1016/j.rmed.2014.10.010.

24. Larsson SC, Gill D. Genetic predisposition to allergic diseases is inversely associated with risk of COVID-19. Allergy 2021 Dec 31, https://doi.org/10.1111/all.14728.

25. Keswani A, Dhana K, Rosenthal JA, Moore D, Mahdavinia M. Ann Allergy Asthma Immunol. 2020;125(4): 479–481, https://doi.org/10.1016/j.anai.2020.07.012.

26. Jackson DJ, Busse WW, Bacharier LB, et al. Association of respiratory allergy, asthma, and expression of the SARS-CoV-2 receptor, ACE2. J Allergy Clin Immunol 2020; https://doi.org/10.1016/j.jaci.2020.04.009.

27. Wan Y et al. Receptor Recognition by the Novel Coronavirus from Wuhan: an Analysis Based on Decade-Long Structural Studies of SARS Coronavirus. J. Virol. 2020. [Epub ahead of print]. https://jvi.asm.org/content/94/7/e00127-20.long

28. Licari A, Votto M, Brambilla I, Castagnoli R, Piccotti E, Olcese R., Tosca MA, Ciprandi G, Marseglia GL. Allergy and asthma in children and adolescents during the COVID outbreak: What we know and how we could prevent allergy and asthma flares. Allergy. 2020 Sep;75(9):2402–2405, https://doi.org/10.1111/all.14369.

29. Lindsley AW, Schwartz JT, Rothenberg ME. Eosinophil responses during COVID-19 infections and coronavirus vaccination. J Allergy Clin Immunol. 2020 Jul;146(1):1–7, https://doi.org/10.1016/j.jaci.2020.04.021.

30. Carli G, Cecchi L, Stebbing J, Parronchi P, Farsi A. Is asthma protective against COVID-19? Allergy 2020;76(3): 866–868, https://doi.org/10.1111/all.14426.

31. Balz K, Kaushik A, Chen M et al. Homologies between SARS-CoV-2 and allergen proteins may direct T cell-mediated heterologous immune responses. Sci Rep 11, 4792 (2021), https://doi.org/10.1038/s41598-021-84320-8.

32. Kapoor R, Sharma B, Kanwar SS. Antiviral Phytochemicals: An Overview. Biochem Physiol. 2017;6:220, https://doi.org/10.4172/2168-9652.1000220.

33. Linnakoski R, Reshamwala D, Veteli P, Cortina-Escribano M, Vanhanen H, Marjomäki V. Antiviral agents from fungi: diversity, mechanisms and potential applications. Front. Microbiol. 2018; 9: 2325, https://doi.org/10.3389/fmicb.2018.02325.

